# Comparison of Push-out Bond Strength in Fiber Posts Cemented with Three Different Cements: Glass Ionomer, Self-Etching, and Self-Adhesive Resin Cement

**DOI:** 10.1101/2025.03.27.25324609

**Authors:** Hossein soraghi, Elnaz Shafigh, Siavash Abdollahpour

## Abstract

**Background and Objective:** This study aimed to compare the push-out bond strength of fiber posts cemented with three different types of cements: glass ionomer, self-etch resin, and self-adhesive resin cement. The objective was to identify which cement provides the most effective bonding in different sections of the root canal.

**Methods:** Thirty extracted central teeth were prepared and divided into three groups, each receiving a different type of cement. The roots underwent standard preparation, including trimming, manual and mechanical filing, and chemical treatment. The push-out test was employed to measure the bond strength of the fiber posts in the coronal, middle, and apical sections of the root canal.

**Results:** The study revealed that in the coronal section, there were no significant differences among the groups. However, in the middle and apical sections, both the self-adhesive and self-etch groups demonstrated a statistically significant increase in bond strength compared to the glass ionomer group.

**Conclusion:** The findings suggest that the choice of cement significantly affects bond strength, particularly in the coronal section of the root canal. Self-adhesive resin cements emerge as a preferred option due to their higher bond strength. This study provides crucial insights for dental practitioners in selecting appropriate cements for different root canal sections, enhancing the longevity and effectiveness of dental restorations.

## Introduction

Root canal-treated teeth, unlike their vital counterparts, frequently require additional means of retention for effective restoration. ^1^This need is particularly acute in teeth where over 50% of the crown structure is lost, necessitating the use of a post within the canal to enhance and secure the retention of restorations ^2^. Notably, the primary function of these posts is not just to physically reinforce the remaining tooth structure, but also to significantly increase retention, a concept supported by numerous studies. ^3^

For many years, the dental industry predominantly relied on cast posts and core restorations for treating root canal teeth. However, the drawbacks of metal posts, such as high root fracture incidence, lack of natural tooth translucency, concerns over corrosion, and the risk of root perforation during post removal, have prompted a reevaluation of these traditional systems [5, 6]. These disadvantages, alongside observations that cast posts may reduce the fracture resistance of the restored tooth, have led to the development of alternative post systems, particularly in cases where mechanical retainers are minimal or absent. In this context, fiber-reinforced composite (FRC) posts have emerged as a popular choice for the restoration of root canal-treated teeth since their introduction in the early 1990s.^4^

FRC posts are predominantly utilized to provide robust support and retention for root canal restorations. Their modulus of elasticity, closely mirroring that of dentin, facilitates favorable stress distribution and minimizes the likelihood of catastrophic fractures, such as root fractures below the bone surface and linear fractures (Kaizer). This unique property allows them to absorb and distribute forces along the root, thereby diminishing the potential for root fractures. Further, their integration with an adhesive bond to root canal dentin and a resin build-up core makes it possible to restore root canal teeth while conserving as much of the remaining tooth structure as possible [8]. The adhesive bond of FRC posts additionally safeguards the sub-tooth structures. Their utility is also evident in the prosthetic reconstruction of wide root canals ^5^.

A significant challenge with FRC posts is the potential for adhesion failure, particularly debonding within the root canals. This issue is often attributed to gaps in the contact surfaces between the post, the cement, and the dentin, a concern noted in several studies ^6^. To mitigate this, dual-cure resin cements are frequently employed, ensuring a seamless adhesion within the root canal. This study delves into the use of both glass ionomer and self-adhesive resin cements for the cementation of FRCs. Introduced in 2002, self-adhesive resin cements have revolutionized the cementation process with their micro-mechanical grip and chemical adhesion capabilities ^7^. These cements contain multifunctional hydrophilic monomers with phosphoric acid groups, enabling reactions with hydroxyapatite, penetration of the smear layer, and creation of porosity. Their chemical interaction with hydroxyapatite ensures a robust adhesion to dentin^8^.

The bonding efficacy of these resin cements is heavily dependent on the quality of the hybrid layer, a factor influenced by several variables including dentin morphology, environmental conditions, working conditions, the chosen bonding system, and the specific properties of the luting cement ^9^. Creating a reliable hybridization in the apical third of the post space is particularly crucial due to the challenges in achieving adhesion in this area. Various cements and adhesive approaches have been explored for effectively connecting FRCs to root dentin ^10^. High bond strength has been observed with dual polymerization resin cements used in conjunction with appropriate dentin preparation techniques (2 or 3-step etch-and-rinse adhesive systems).

Self-adhesive resin cements represent a recent advancement in this field. According to manufacturers, these cements eliminate the need for pretreatment of the tooth surface, integrating the etching and priming stages into a single step. This integration significantly reduces technical sensitivity and enhances the procedural simplicity. Glass ionomer cements (GIC) and resin-modified GICs (RMGICs) have also been investigated for their efficacy in bonding FRCs ^11^. A notable feature of GICs and RMGICs is their hygroscopic expansion post-water absorption, which compensates for their initial shrinkage ^12^. This characteristic suggests that the residual water in dentinal tubules might aid the hygroscopic expansion of GICs and RMGICs in FRC bonding. The resistance to post fiber displacement is directly linked to their adhesion to root dentin ^13^.

The enduring success of root canal treatments heavily relies on the effectiveness of post-cementation techniques. With the evolution of dental materials and techniques, the choice of an optimal cement for securing fiber posts has become a focal point in endodontic research. Variations in the properties of different cements, such as adhesion strength, ease of application, and compatibility with dental tissues, can significantly impact the longevity and reliability of post-endodontic restorations. Furthermore, the interaction between these cements and the unique anatomical and structural features of root canal-treated teeth presents a complex challenge, necessitating a deeper understanding of their bonding mechanisms. Recent advancements in dental materials, particularly in resin-based cements, have introduced new possibilities for enhancing the quality of post-cementation ^14^. However, there remains a gap in comprehensive comparative studies that rigorously examine the performance of these materials in different root canal regions. This gap highlights the need for systematic research to guide clinicians in selecting the most suitable cement based on specific clinical scenarios and root canal characteristics. Addressing this need, our study aims to provide a detailed comparison of the bond strengths of widely used cements, thereby offering valuable insights into their application in different root canal sections. Therefore, the primary aim of this study is to evaluate and compare the push-out bond strength of glass ionomer cements, resin cements, and self-adhesive resin cements in fiber posts. ^15^

## Materials and Methods

### Study design

This study investigated the push-out bond strength of fiber posts cemented with three different types of cements: self-adhesive resin, self-etch resin, and glass ionomer. The methodology encompassed the collection and preparation of thirty extracted teeth, their division into three groups for distinct cementation processes, and subsequent analysis. Each group underwent a specific protocol for root canal preparation, cement application, and post cementation. The bond strength was evaluated using a push-out test, and the results were statistically analyzed to compare the performance of the various cements. This in vitro research, conducted in compliance with ethical standards, aimed to enhance the understanding of effective cementation techniques in dental restoration.

### Sample Collection and Preparation

Thirty central teeth, extracted due to periodontal issues, were selected based on similar anatomical structures and absence of structural defects or caries. This in vitro study was approved by the Research Ethics Committee of the School of Dentistry at Aja University of Medical Sciences, Iran (Ethical Code: IR. AJAUMS.REC1400.170). Consent of participate was deemed unnecessary recording to national regulation. The roots were uniformly trimmed to a length of 16 mm, measured from the apex to the cervical area.

The teeth underwent endodontic treatment as follows: We used a ProTaper Sx rotary device (Dentsply Miallefer, Ballaigues, Switzerland) to enlarge the orifice and the canal’s coronal third.

The irrigation used in this initial step was only normal saline. Using a #10 K file, we established that the working length was 1 mm short of the apical foramen. We prepared the roots with the rotary device up to #F3 in association with irrigation solutions. The standardized irrigation method was performed for teeth using a 5 mL disposable plastic syringe and a 31 gauge side vented needle, the quantity and contact time of each solution were standardized to 5 mL of 5.25% NaOCl (ChloraXiD, PPH Cerkamed, Stalowa Wola, Poland) for 1 min after each file, then 5 mL of 17% EDTA (Pulpdent, Watertown, MA, USA) for 2 min, and finally 5 mL of 5.25% NaOCl for 1 min to remove the smear layer (Özyürek et al. 2018). Canal preparation was done chemomechanically using the step-back technique with MAF 35 and a size 35 MAC for all samples. Canal filling was done using lateral condensation with gutta-percha and AH 26 sealer. After the sealer setting was completed, the post space in each sample was prepared to a length of 10 mm using a size 2 fiber post drill. (Figure 1). ^16^

**Figure 1.**
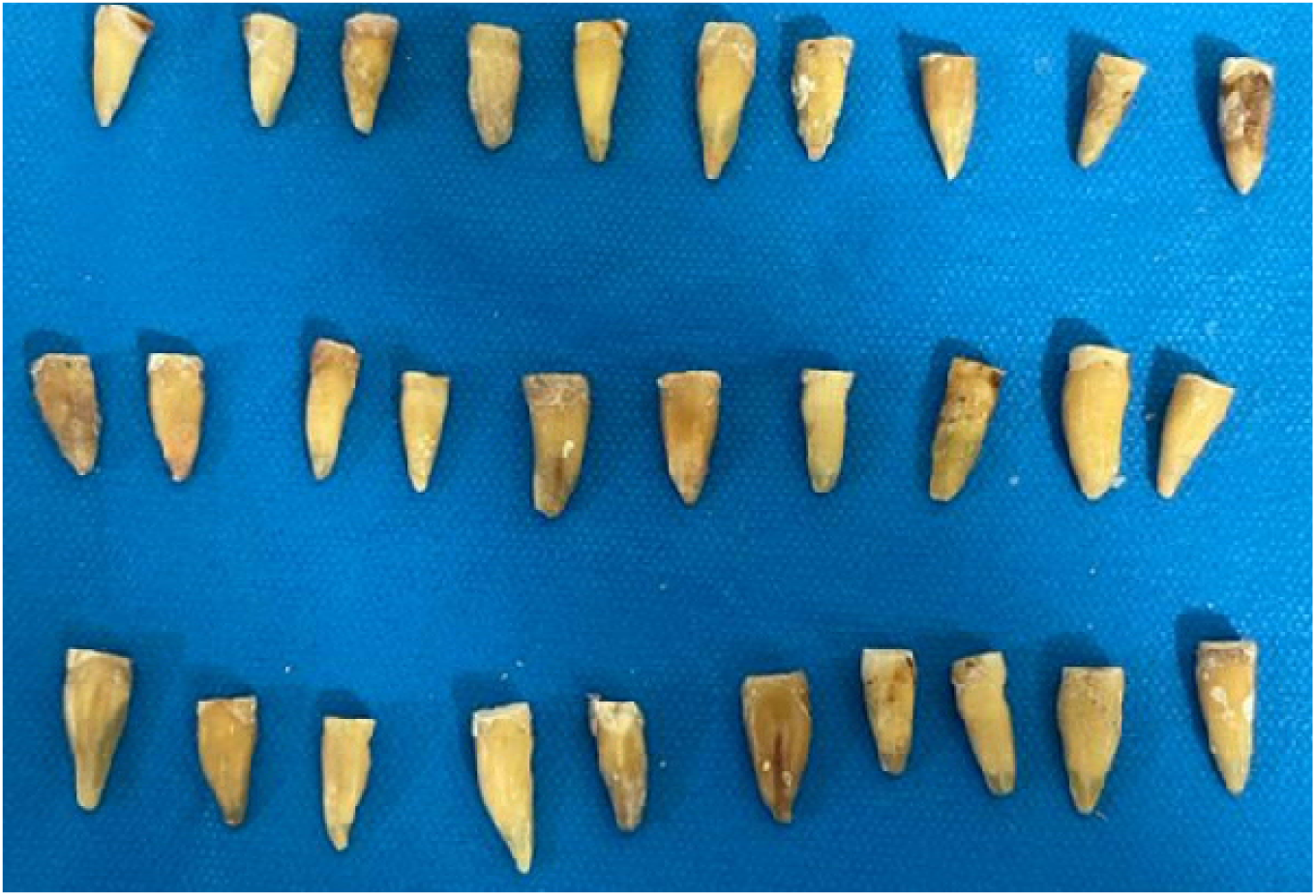
Teeth after preparation

### Samples

The teeth were randomly divided into three groups of ten, each assigned to a different cementation system (Figure 2).

**Figure 2.**
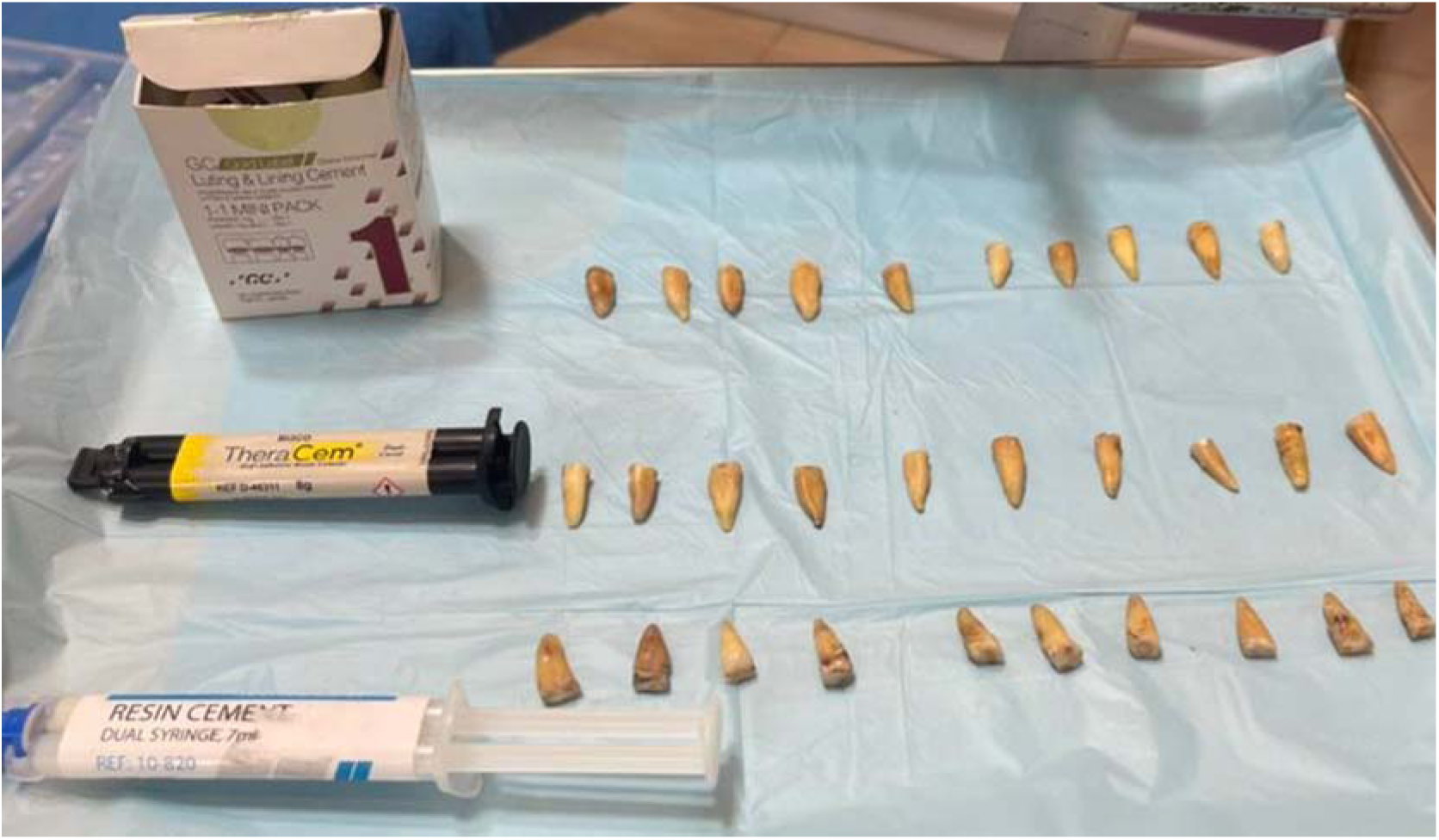
Teeth before cementing

**Group 1 - Self-Adhesive Resin Cement (Bisco, Theracem, USA):** Root canals were etched with 37% phosphoric acid for 15 seconds, rinsed, and dried. The post surfaces were cleaned with 95% ethanol, etched with phosphoric acid for 60 seconds, rinsed, and dried. Cement was then manually inserted into the canal using a lentulo spiral.

**Group 2 - Self-Etch Resin Cement (Masterdent, China):** The post surfaces were cleaned with 95% ethanol. The self-etch resin cement was then manually placed into the root canal using a lentulo spiral.

**Group 3 - Glass Ionomer Cement (GC, Japan):** Post surfaces were cleaned with 95% ethanol. After mixing the glass ionomer according to the manufacturer’s instructions, it was manually placed in the canal with a lentulo spiral. (table 1)

**Table 1.** Composition of cements used in the study.

| Type of cement | Theracem | Masterdent | Glass Ionomer |
| --- | --- | --- | --- |
| Curing type | Self-adhesive | Self-etch | Self-etch |
| Compositions | Base: Calcium filler, glass filler, dimethacrylates, ytterbium fluoride, initiator, amorphous silica<br>Catalyst: Glass filler, methacryloyloxydecyl dihydrogen-phosphate (MDP), amorphous silica | Base: Methacrylate monomers, alkaline fillers, silanated fillers, initiator, pigments<br>Catalyst: methacrylate monomers containing phosphoric acid groups, initiator, stabilizer | Powder<br>Fluoroaluminosilicate glass<br>Liquid<br>Poly acrylic acid |

The composition of cements is shown in table 1.

### Post Cementation and Sectioning

In all groups, after cementing the post at the specified length, excess cement was removed with a micro brush. samples according to the above methods were washed for 10 seconds and then dried with compressed air. All samples were mounted in cylindrical plastic molds in transparent acrylic. After 24 hours, cross-sectional samples were obtained from the root area containing the post using a 0.20 mm thick diamond disk sections (Hager and Meisinger GmbH, Neuss, Germany), at a speed of 150 rpm under water resulting in two 4 mm samples. To evaluate the adhesion of the posts, each root was sliced into two -. The sectioning started 1 mm below the CEJ: there was a coronal (B), and 7mm under the CEJ was an apical (A) section with a thickness of 4 mm each. ^17^The samples were restored in 37^□^ distilled water until the test which was 2 weeks long.^18^ (figure 3)

**Figure 3.**
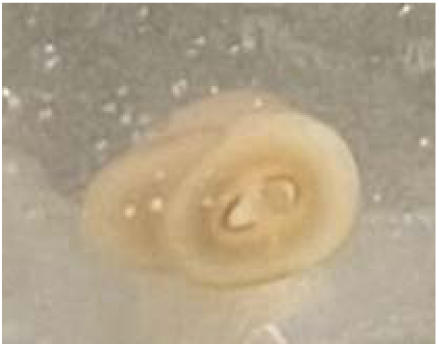
Cut sections

### Push-Out Test

The prepared samples underwent a push-out test using an electromechanical testing device (Zwick/Rowell Z020, Switzerland), with the following parameters: 2.5 mm/min crosshead speed, 5 kN Instron^®^. The forces as a function of the displacement of fiber posts were recorded determined the peak force. The test proceeded until complete separation of the fiber post from the root canal, and the bond strength was measured in MPa (Figure 4). which was calculated using the following formula for each sample, Failure was identified by observing the curve drop drawn by the Instron software. The applied force increased gradually, and the device recorded the maximum force at the moment of failure. ^19^

**Figure 4.**
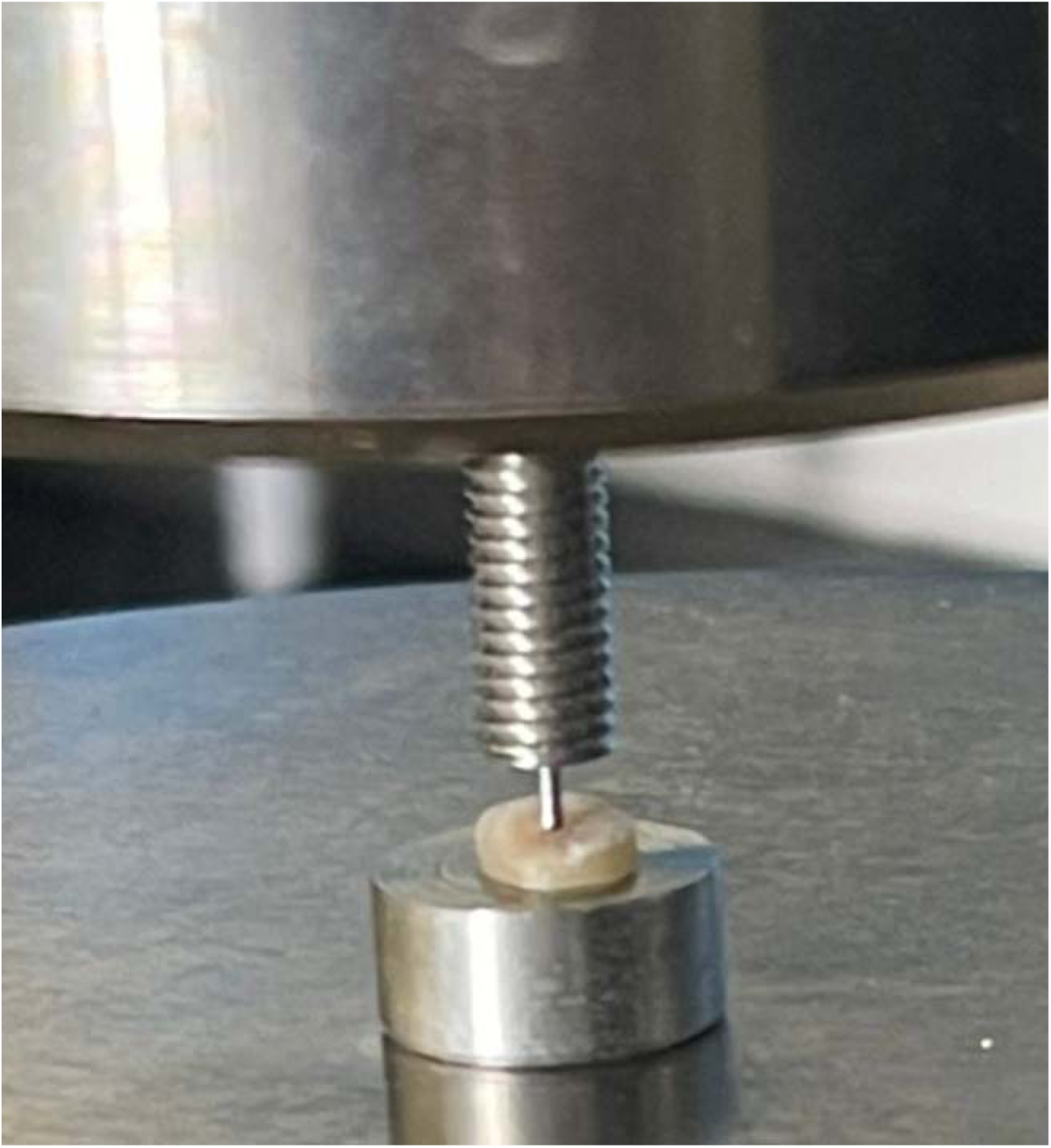
During Push-out test

### Statistical Analysis

### Data Analysis

The mean and standard deviation were calculated, and data were analyzed. The normal distribution of the data was determined using the Kolmogorov-Smirnov test in SPSS software. version 23.0; SPSS, Chicago, IL, USA) was used. The Kolmogorov-Smirnov test and Shapiro-Wilk test were used to check the distribution of the data. We used one-way ANOVA for bond strength comparison among the six groups and Dunnett’s multiple comparison t-test for pairwise group comparisons

## Results

The study’s findings revealed distinct patterns in the push-out bond strengths among the three groups of cemented fiber posts. Notably, there was no significant difference in the push-out bond strength of the coronal sections across the three groups. However, in the middle and apical sections, both the self-adhesive and self-etch groups demonstrated a statistically significant increase in bond strength compared to the glass ionomer group. Despite the self-adhesive group exhibiting higher bond strength values than the self-etch group in these sections, the differences were not statistically significant (Figure 5).

**Figure 5.**
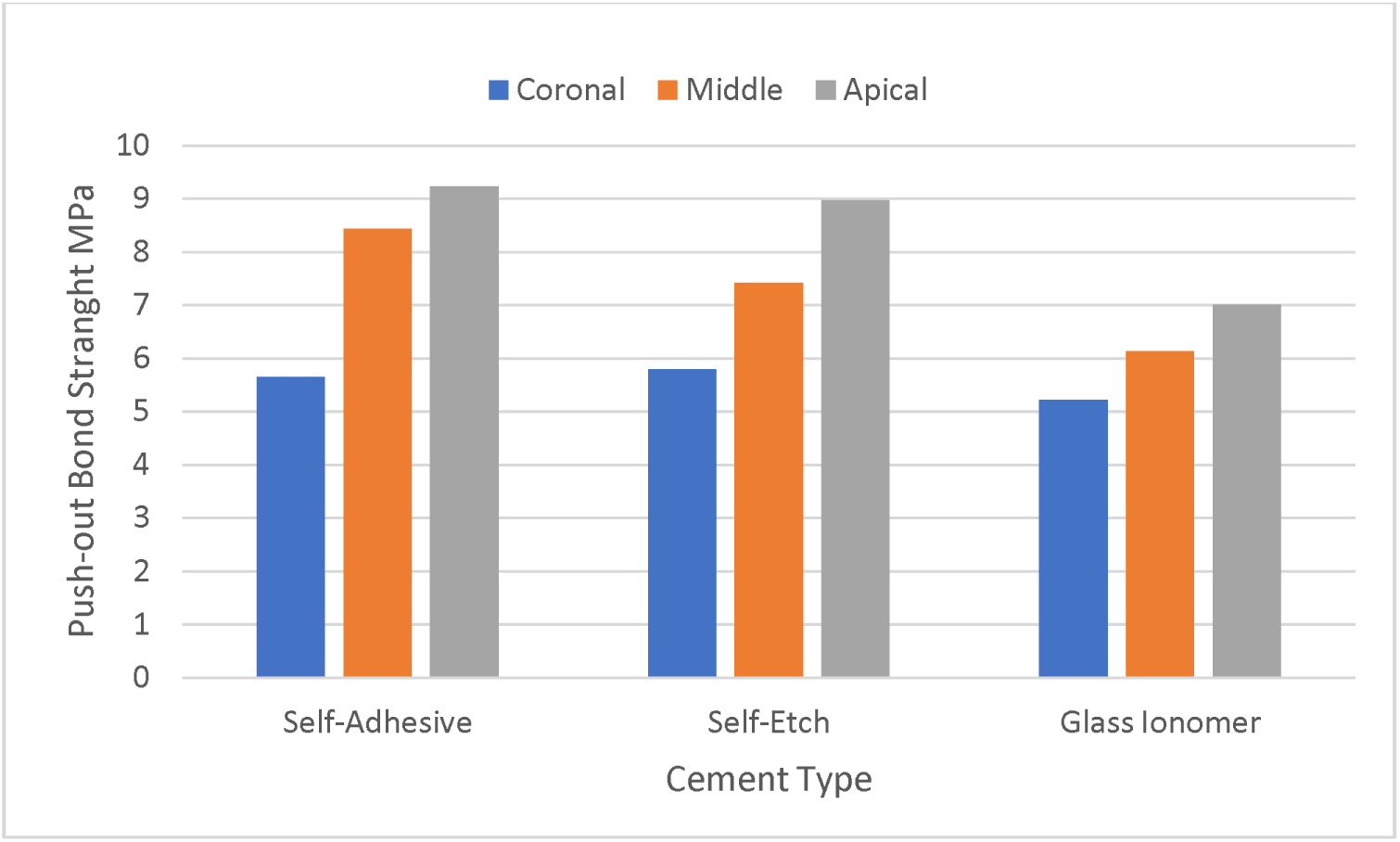
Comparison of push-out bond strength values between three types of self-adhesive, self-etch and glass ionomer cements in three coronal, middle and apical sections

A consistent trend observed across all groups was the variation in bond strength across different sections of the root canal. The push-out bond strength values in the apical sections were significantly higher than those in the middle sections. Similarly, the bond strengths in the middle sections showed a significant increase compared to the coronal sections (Figure 5).

These results indicate that while the type of cement affects the bond strength in different sections of the root canal, the location within the canal (coronal, middle, or apical) also plays a critical role in determining the effectiveness of the bonding (Figure 5).

The mode of failure in each group is shown in figure 6. In all groups debonding in cement-dentin interface was higher and no statistical difference was observed between groups.

**Figure 6.**
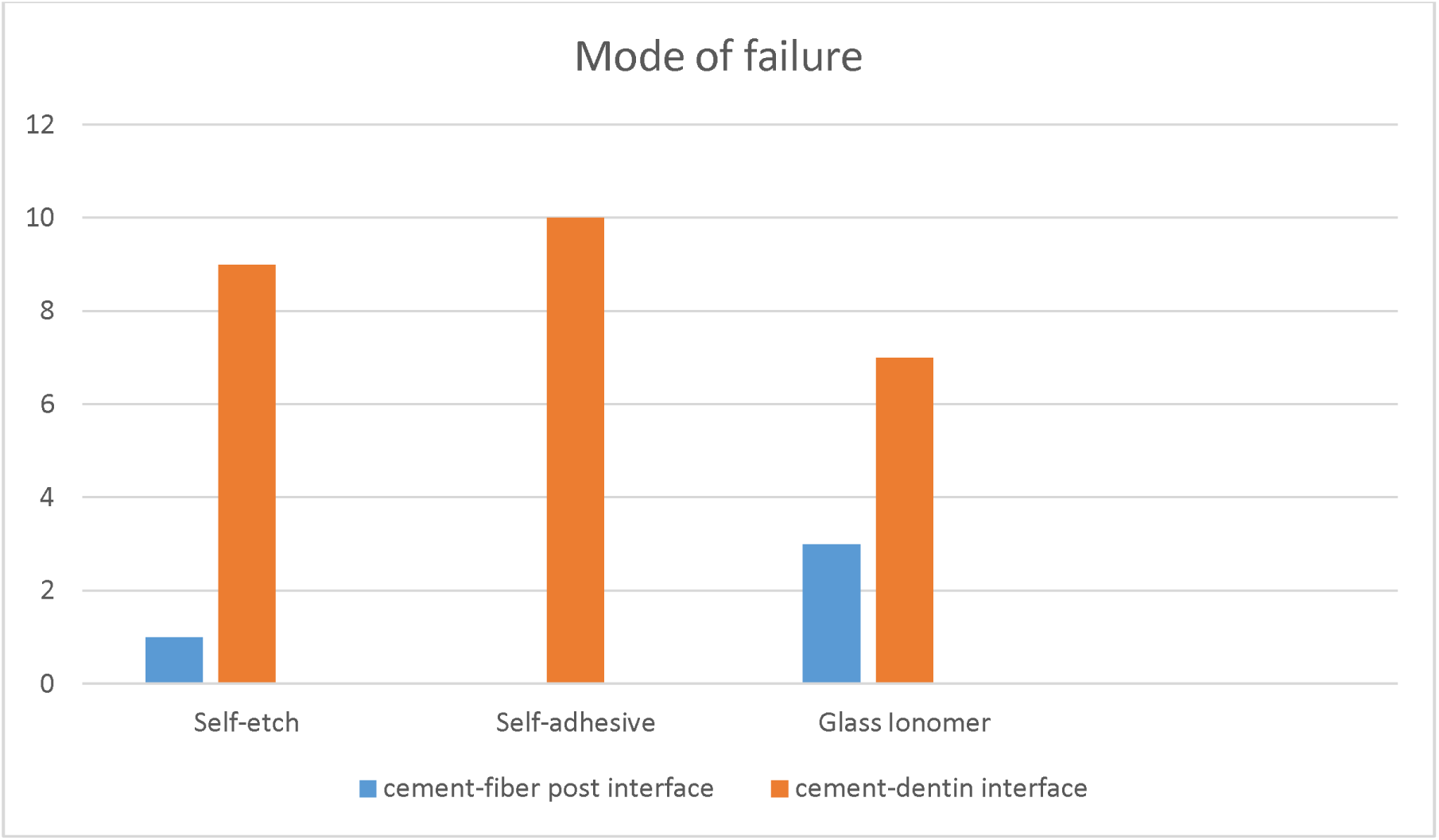
Mode of failure in each group

## Discussion

Our study aimed to compare the push-out bond strength among three conventional cements: glass ionomer, self-etch resin, and self-adhesive resin cement in the context of cementing fiber posts. Because of a little evidence to discuss a better bond between self-etch and self-adhesive cements and because of many importance of correct usage of cements in sealing the root materials we think that this article can be helpful in better understanding and better employments of these materials.

The results revealed a nuanced pattern: while the coronal section exhibited a significant increase in bond strength for both self-adhesive and self-etch groups over glass ionomer, the middle and apical sections did not display any significant differences among the groups. This suggests a differentiated impact of cement types depending on the root canal section, underscoring the importance of cement selection in different canal regions.

The push-out test was the chosen methodology for its closer simulation of clinical conditions compared to linear shear tests, effectively replicating the shear stress at the post-cement and cement-dentin interfaces ^20^. Its selection has been validated by studies like that of Goracci et al. (2004), which underscored its efficiency and reliability in measuring the bond strength of fiber posts to root canal dentin ^21^.

Bond strength to dentin is a complex interplay of various factors, including dentin morphology, polymerization methods, compatibility between resin cement and bonding agent, as well as the type of washing solutions and sealers used in root canal treatment. The method of adhesive application also significantly impacts this dynamic ^22^. Our findings indicated that self-adhesive cements, particularly Theracem, exhibited higher bond strength. This could be attributed to their unique adhesion mechanism, involving both micromechanical engagement and chemical reactions between the acidic monomer groups and hydroxyapatite, chelating calcium ions and enhancing adhesive chemical adhesion ^23^.

The glass ionomer cement forms its primary bond to the dentin substrate through a chemical interaction between the carboxylate groups and calcium hydroxyapatite during the acid-base reaction. Comparatively, self-adhesive resin cement, while sharing some similarities in chemical reactions, benefits from an additional micro-mechanical bond, potentially accounting for its observed increase in bond strength over glass ionomer.

Water immersion and its effect on cement bond strength were also considered in our study, given the critical role of humidity in root canal treatments. Consistent with the findings of previous studies, we observed that self-adhesive cement’s bond strength can increase after 24 hours of water immersion, highlighting environmental conditions’ influence on bonding efficacy [26].

However, the challenge of controlling humidity levels in root canals, compounded by limited visibility and moisture retention due to surface tension, is a notable concern. This issue is particularly problematic for self-etch systems that are typically applied on dry dentin ^24^. Therefore, the increased humidity within the root canal could potentially decrease the bond strength of self-etch systems, even with careful drying efforts.

The study also revealed higher bond strength in the apical region, especially with self-adhesive resin cements, suggesting a reduced sensitivity to variations in dentin depth and tubule density compared to other cements. This is in line with the findings of Lo Giudice et al. (2015), who reported variations in tubule density across different root canal regions ^25^.

Despite these insights, some limitations of self-adhesive cements were noted. Electron microscope examinations have shown that these cements do not always achieve complete demineralization of collagen, resin penetration into the dentin, and thorough removal of the smear layer, which could impact the bond strength with dentin. In contrast, the bond strength in self-etching cements appears to be more influenced by the dentin area than the density of dentinal tubules ^26^. Furthermore, our results align with those of other studies, such as those, who reported higher bond strength values in the apical third than in other parts of the root canal. This could be indicative of the inherent differences in the structural composition of dentin across various canal regions ^27^, which in turn affects the bonding efficacy of different cements.

In examining the broader landscape of research in this area, it becomes evident that the findings on the comparative effectiveness of self-etch and self-adhesive resin cements have been mixed. While some studies report weaker bonds with dentin for self-adhesive resin cements, others find no significant difference in bond strength when different resin cements are used for cementing fiber posts to dentin ^28^. This variability in results underscores the complexity of factors influencing bond strength, including the type of resin cement, the specific application method, and the inherent properties of the dentin substrate ^29^.

Our study contributes to this ongoing discourse by offering specific insights into the performance of three widely used cements in dental practice. The findings suggest that while each cement type has its inherent advantages and limitations, the choice of cement should be guided by the specific requirements of the root canal section being treated. This approach can help optimize the longevity and efficacy of dental restorations, ensuring better outcomes for patients. In conclusion, our study not only sheds light on the comparative bond strengths of different cements but also underscores the importance of considering the unique characteristics of each root canal section in the selection and application of cements for fiber post cementation. By doing so, dental practitioners can make informed decisions that enhance the quality and durability of dental restorations, ultimately contributing to better oral health outcomes.

## Conclusion

Our study extensively compared the push-out bond strength of fiber posts using three types of cements: glass ionomer, self-etch resin, and self-adhesive resin cement. The results revealed nuanced differences in bond strength across different sections of the root canal, highlighting the importance of cement selection in dental restoration. Specifically, in the middle and apical, both self-adhesive and self-etch cements showed significantly higher bond strength compared to glass ionomer, though no significant differences were observed in the coronal sections. The choice of the push-out test as the evaluation method was validated by its effectiveness in simulating clinical conditions and its reliability in measuring bond strength.This insight is crucial for dental practitioners when choosing cements for different root canal sections. The study also highlighted the impact of environmental factors such as humidity on the bonding efficacy of cements. Controlling moisture levels in root canals is challenging but crucial, as it significantly influences the bond strength, especially in self-etch systems.

Our results suggest that the bond strength in the apical region is generally higher, a finding that is consistent with other research in the field. However, the limitations of self-adhesive cements, such as incomplete demineralization of collagen and inadequate resin penetration, point towards the need for careful consideration in their application. In conclusion, our study underscores the complexity of factors influencing the bond strength of different cements in root canal-treated teeth. It provides valuable insights for dental practitioners in selecting the appropriate cement based on the specific requirements of the root canal section. Ultimately, this research contributes to enhancing the quality and durability of dental restorations, improving patient outcomes in dental health care.

## Declarations

## Consent for publication

Not applicable

## Data availability statement

The data that support the findings of this study are available from AJA University of Medical Sciences. Restrictions apply to the availability of these data, which were used under license for this study. Data are available from Dr. ELNAZ SHAFIGH with the permission of AJA University of Medical Sciences.

## Data Availability

All data produced in the present study are available upon reasonable request to the authors

## Acknowledgement

The authors would like to thank the members of AJA University of Medical Sciences for their unwavering support.

## Ethical Considerations

This in vitro study was approved by the Research Ethics Committee of the School of Dentistry at Aja University of Medical Sciences, Iran (Ethical Code: IR. AJAUMS.REC1400.170). Consent of participate was deemed unnecessary recording to national regulation.

---

**Table 1:**
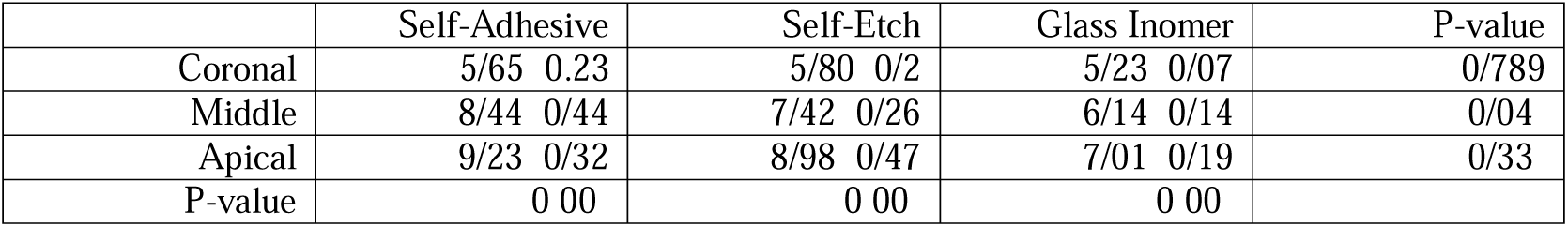
Comparison of push-out bond strength values between three types of self-adhesive, self-etch and glass ionomer cements in three coronal, middle and apical sections.

**Table 2:**
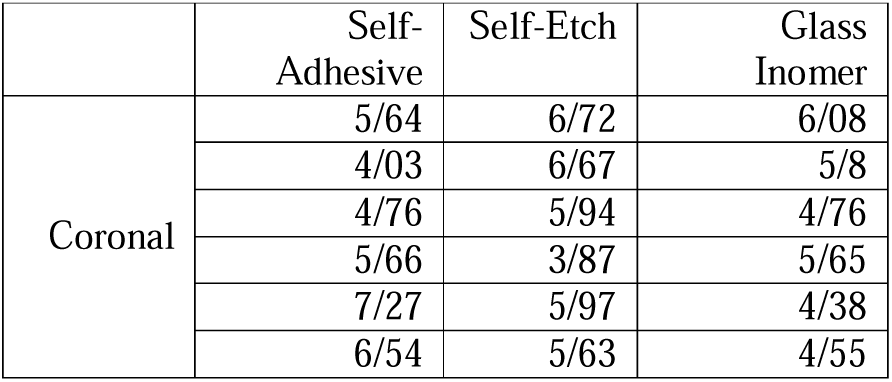
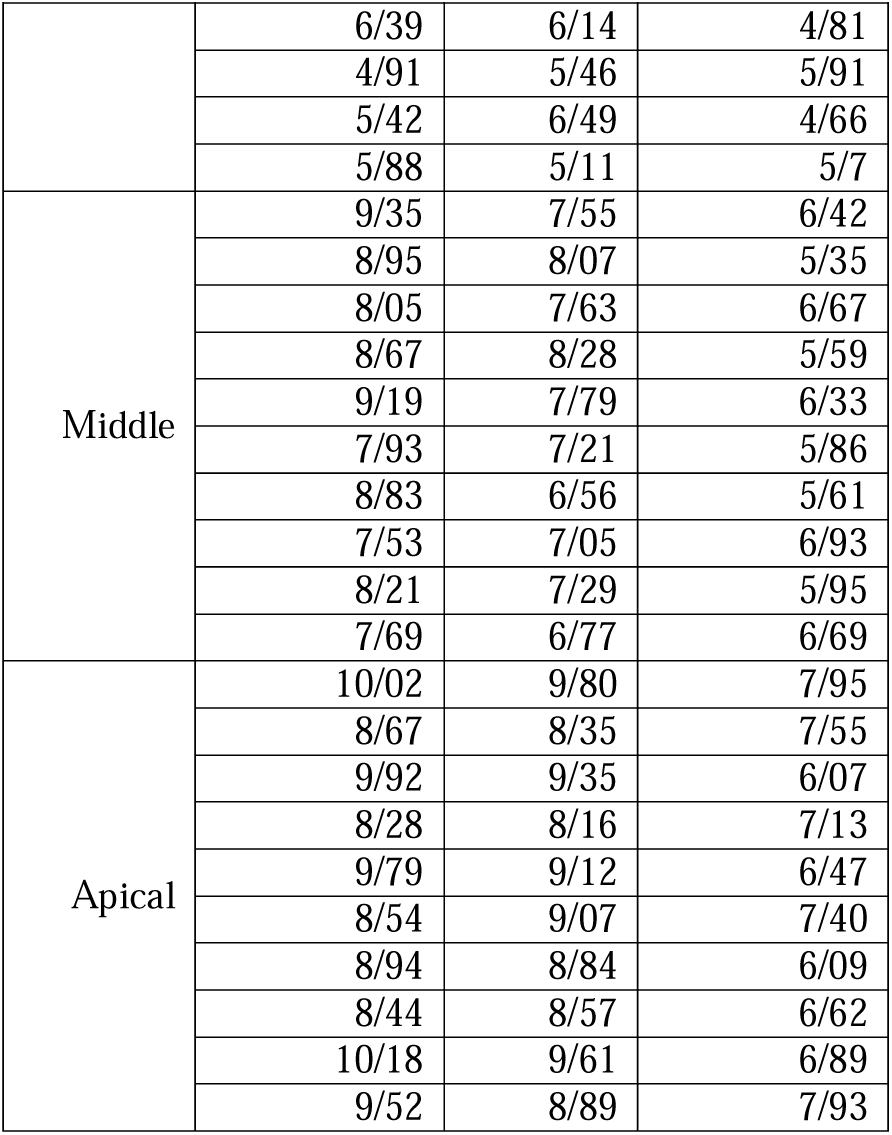
Push-out bond strength of all studied samples in MPa.

**Diagram 1:**
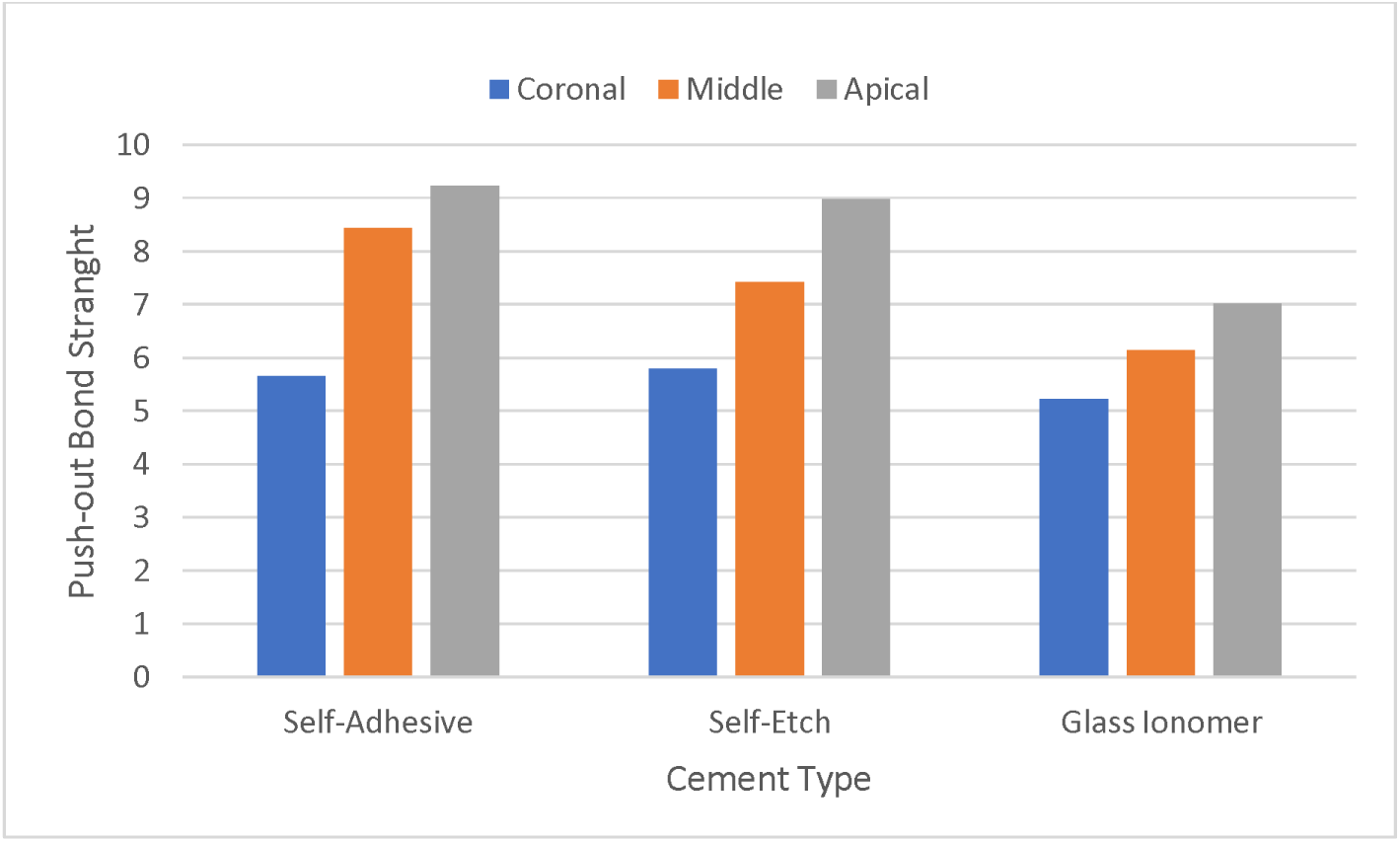
Comparison of push-out bond strength values between three types of self-adhesive, self-etch and glass ionomer cements in three coronal, middle and apical sections

